# Time to death and its predictors among neonates with perinatal asphyxia at a tertiary hospital in southern Ethiopia

**DOI:** 10.1101/2024.08.13.24311935

**Authors:** Jenenu Getu Bekele, Niguse Mekonnen Kara, Amene Abebe Kerbo, Tadiwos Utalo Urkashe

**Affiliations:** Department of Epidemiology and Biostatistics, College of Health Sciences and Medicine, Wolaita Sodo University, Sodo, Ethiopia; Department of Reproductive Health, College of Health Sciences and Medicine, Wolaita Sodo University, Sodo, Ethiopia; School of Public Health, College of Health Sciences and Medicine, Wolaita Sodo University, Sodo, Ethiopia

## Abstract

**Background:** Perinatal asphyxia (PNA) remains a leading, yet preventable, cause of neonatal death, disproportionately affecting low-resource settings like Ethiopia. Despite its significance, data on the burden and factors influencing survival among asphyxiated neonates in this region are scarce. This study aims to address this gap by investigating the time to death and its associated predictors in this vulnerable population.

**Methods:** A retrospective cohort study was conducted at Wolaita Sodo University Comprehensive Specialized Hospital’s Neonatal Intensive Care Unit (NICU) in southern Ethiopia. Medical records of 404 neonates diagnosed with PNA between January 2019 and December 2023 were reviewed. The study followed these neonates for a total of 2889 person-days to assess their survival outcomes. Kaplan-Meier analysis estimated the median time to death, and a Weibull regression model identified independent predictors of mortality.

**Results:** The findings revealed the incidence density of PNA-related mortality of 30.8 per 1,000 person-days (95% CI: 25.0-37.9). Notably, nearly 72% of deaths occurred within the first critical week of life, with a median survival time of 20 days. The multivariable Weibull regression analysis identified several factors significantly associated with shorter time to death at p 0.05. These are presence of meconium-stained amniotic fluid (MSAF), low fifth-minute Apgar score (less than 7), birth weight greater than 4000gm, low admission oxygen saturation level, treatment with anticonvulsant, the use of Continuous Positive Airway Pressure (CPAP) for oxygen administration, and need for resuscitation with chest compression.

**Conclusion:** This study highlights the significant burden of PNA-related mortality, particularly during the first week of life, in a resource-limited setting. The findings underscore the urgent need for improved PNA management strategies. Furthermore, specific treatment decisions, including the use of anticonvulsants, oxygen therapy methods, and resuscitation techniques, emerged as crucial factors influencing survival outcomes. These results call for further investigation into these specific interventions and potentially revising PNA management protocols to optimize the chances of survival for asphyxiated neonates in low-resource settings like Ethiopia.

## Introduction

Perinatal asphyxia (PNA) is a public health problem and it is responsible for 30%–35% of neonatal deaths worldwide. The World Health Organization (WHO) defines PNA as the failure to initiate and sustain breathing at birth. It can happen when the intrauterine (placental gas exchange) or immediately after birth (pulmonary gas exchange) is compromised or ceased all together. This causes partial or complete lack of oxygen to the vital organs of the neonate which eventually leads to asphyxia. Neonatal hypoxic-ischemic encephalopathy (HIE) refers to the neurologic consequences of perinatal asphyxia which is an important cause of permanent damage to CNS cells which intern results in neonatal death, cerebral palsy, and mental deficiency [1–5].

The majority cases of PNA occur during the labor and delivery process, although up to 20% can occur during pregnancy and in the early post-natal period. It may result from different causes such as; maternal hemodynamic compromise due to hemorrhage, amniotic fluid embolism; placental events (acute abruption), uterine events (rupture), cord events (tight nuchal cord, cord prolapse) and intrapartum infection (chorioamnionitis). Therefore, early diagnosis and prompt management of these complications in the delivery room is a crucial step in the prevention of late sequels of PNA [6–8].

Neonatal period which is the first 28 days of life is among the most vulnerable periods of life and requires intensified quality intrapartum and newborn care. Globally 2.4 million children died in this period in 2020. There are approximately 6700 newborn deaths every day, which is equal to 47% of all child deaths under the age of 5 years. To avert this tragic loss of neonates, the United Nations (UN) through its Sustainable Development Goal (SDG) goal-three aims to reduce neonatal mortality to at least as low as 12 deaths per 1,000 live births by 2030. But many developing countries seem far from achieving the target [8–10].

Globally, PNA accounts for an estimated 900,000 deaths each year and is one of the primary causes of early neonatal mortality. Of those infants affected by PNA, 15-20% dies in the neonatal period, and up to 25% of the survivors are left with permanent neurologic deficits. Among the survivors of severe PNA, majority will leave with some form of handicap such as cerebral palsy, developmental delay, visual and hearing impairment, and learning and behavioral problems. Therefore, apart from studying neonatal mortality due to PNA, it is important to study the survival status and short and long term effects of the condition [2, 11, 12].

The exact magnitude of perinatal asphyxia in low and middle-income countries is not known since more than 60% of births take place outside facilities. But it is evident that intrapartum related neonatal death (birth asphyxia) is a leading cause of neonatal mortality in low-income countries largely due to poverty, large family size, and cultural beliefs. Neonatal mortality is an indication of the economic status of a country. The incidence of perinatal asphyxia is 2/ 1000 births in developed countries, but the rate is up to 10 times higher in developing countries where there is limited access to maternal and neonatal care services [8, 13, 14].

In 2021, Ethiopia was ranked fourth among the top ten countries in the world with the highest neonatal mortality, having 97,000 neonatal deaths with a neonatal mortality rate of 33 deaths per 1000 live births (LBs). The commonest causes of neonatal deaths are preterm birth, perinatal asphyxia, and infections. Ethiopia’s health sector transformation plan II has set a goal of lowering the neonatal mortality rate to 21 per 1000 LBs by 2025 from the current level. PNA is one of the leading causes of neonatal mortality in Ethiopia which accounts for 34% of neonatal mortality [9, 15–17]

While PNA is a recognized issue, comprehensive data on its impact in Ethiopia at a national level remains elusive. Existing studies primarily focus on specific healthcare facilities, resulting in a wide range of estimated PNA-related deaths. For example, the incidence of mortality due to asphyxia is 29, 53, and 100 per 1000 person days in Jimma Medical Center, Dessie Comprehensive Hospital, and public hospitals of Addis Ababa respectively but, it is only 9.1 deaths per 1000person days in health facilities found in South West Ethiopia (18-21). There is also a disparity among the studies done in Ethiopia on the median survival time after perinatal asphyxia. According to a prospective cohort study done in Southern Ethiopia, the median survival time was 6.5 days. Which is much lower as compared with studies done in Jimma, Addis Ababa, and Dessie, where the median survival time is 20, 10, and 8 days respectively (18-20, 22). According to most studies predictors of survival status of asphyxiated neonates can be classified into sociodemographic, obstetric, neonatal characteristics, clinical, laboratory, and management predictors [18–22].

Despite different initiatives and strategies that have been implemented to prevent neonatal death, it is still high and not reduced as expected in developing countries including Ethiopia. Although several studies were made in Ethiopia on neonatal mortality in general and perinatal asphyxia in particular, most focus on rate, and little has been done on the time to death and its predictors.

Identifying local risk factors and their influence on time to death can inform targeted interventions and improve overall neonatal survival rates in Ethiopia. Therefore, this study aims to address the knowledge gap by investigating the following study questions: What is the incidence of neonatal mortality due to perinatal asphyxia at WSUCSH? What is the time to death among asphyxiated neonates admitted to WSUCSH? What are the clinical and sociodemographic factors associated with time to death in this population? By addressing these questions, this study will contribute valuable insights into the local context of perinatal asphyxia and neonatal mortality in Southern Ethiopia. The findings can guide healthcare professionals in identifying high-risk neonates and implementing appropriate interventions to improve their chances of survival.

## Methods

### Study setting, period, and design

The study was conducted in Wolaita Sodo University Comprehensive specialized Hospital (WSUCSH). The hospital is found in Wolaita Sodo Town which is found 360km far from Addis Ababa in the South-West direction. The hospital serves over 2 million catchment population and over 90,000 people visit the outpatient department per year. Neonatal Intensive Care Unit (NICU) is a separate unit in the pediatrics department that started service in December 2014 and it has 30 beds. According to last year’s facility report there was a total of 1444 neonatal admissions out of which 136 of them were PNA cases. One hundred fifty-four neonatal deaths were reported in the same year due to all causes among the total NICU admissions (unpublished data found from the department).

Institutional based retrospective cohort study was conducted. We reviewed the medical records of patients who were admitted to the NICU of WSUCSH over a five years period from January 2019-December 2023. The data collection was carried out from January 01-31, 2024.

### Study population

The source population was all neonates who were admitted with the diagnosis of PNA from January 2019-December 2023 and the study population was all randomly selected medical records of PNA patients who were admitted to the NICU of WSUCSH. Records of neonates whose admission date, discharge date, and outcome were not recorded on the chart and those who had major congenital anomalies were excluded from the study.

### Sample size determination and sampling technique

The sample size was calculated to check the adequacy of the sample size used for a survival sample size calculation power approach with Cox proportional hazard assumption. By using the Schoenfeld formula [23] and considering the following assumptions: Significance level (α) = 0.05, Power 80% (β) = 0.8, proportion of variability among covariates of interest (SD) = 0.5, Square of correlation of independent variables with time to death (R^2^) = 0.5 and by taking cord prolapse, vaginal mode of delivery, stage-III HIE, sepsis, and AKI as the common predictive variables [20,22,24,25], the sample size was calculated using Stata version 14.2 statistical software as shown in **Table 1**.

**Table 1.** Sample size calculation for the study of the predictors of the time to death among PNA patients who were admitted to NICU of WSUCSH; 2024.

| Variables | Probability of events | Probability of withdrawals | Effect size (CHR) | Sample size (n) | Author &(study area) |
| --- | --- | --- | --- | --- | --- |
| Cord prolapse | 7.85% | 10% | 13.68 | 130 | Dessu et al. (Southern Eth) [22] |
| Stage-III HIE | 32.79 | 10% | 2.06 | **408** | Daka et al. (West Oromia) [24] |
| Acute kidney injury | 22.52% | 10% | 2.95 | 265 | Lencho K. et al. (Jimma) [20] |
| Vaginal mode of delivery | 54.08% | 5.9% | 2.91 | 109 | Kebede BF et al. (SW-Eth) [25] |
| Sepsis | 54.08% | 5.9% | 0.48 | 230 | Kebede BF et al. (SW-Eth) [25] |

### Study variables

**The dependent variable** is time to death among neonates who were admitted with the diagnosis of PNA.

**The independent variables include;** sociodemographic and obstetric characteristics of the mothers of PNA patients (age, place of residence, parity, duration of labor, duration of rupture of membrane (ROM), place of delivery, mode of delivery, obstetric complication, etc.), neonatal characteristics (Sex of the baby, gestational age, birth weight and age at presentation), clinical and laboratory variables (Apgar score, crying, stage of HIE, vital signs, baseline lab results), treatment related variables (bag &mask ventilation, chest compression, adrenalin, aminophylline, fluid and electrolyte management, oxygen, anticonvulsant, antibiotics, and calcium gluconate administration.

### Operational definition

**Event:** in this research the event of interest is death due to PNA

**Censored:** are those neonates who are:

- Discharged improved (has discharge summery record),
- Left against medical advice (caregivers signature, or clearly stated on the discharge summery),
- Those referred to other facilities (has copy of referral form in the medical record)
- Those still on treatment after 28 days of follow-up.

**Death:** neonates who were admitted with a diagnosis of perinatal asphyxia and died from any immediate cause secondary to asphyxia while in the NICU of the hospital, for which a death summary sheet was found.

**Time to death:** it is a time during which asphyxiated neonates developed the event of interest measured in days. It is the period from the date of diagnosis of PNA to the date of death.

**Perinatal asphyxia:** was considered based on the clinical diagnosis made by a health care provider and the stage of PNA is also taken as recorded on the medical record of the patient.

**Follow-up period:** The starting point for retrospective follow-up was the time of diagnosis of PNA and the end point was date of death, discharge or 28 days of age after birth. Therefore, the overall follow up period was 28 days.

### Data collection tool and procedure

Data was collected using a data abstraction checklist adapted after reviewing different literature, previous related studies [18–25], the Health Management Information System (HMIS) registration book, standard NICU referral paper, and PNA follow up chart. The checklist contains four parts; questions related to socio-demographic and obstetric related factors, neonatal related factors, clinical and laboratory factors, and management related factors. (See **S3 File** for data abstraction checklist)

### Data processing and analysis

The collected data using the Kobo collect mobile application was submitted to the central computer database and checked for completeness and consistency daily. At the end of the data collection period, the data was exported using excel and SPSS-label syntax command. The data carefully cleaned and coded. Finally, it was exported to Stata version 14.2 (Stata Corp. LP, College Station, Texas) statistical software for analysis.

Incidence density rate of mortality was computed by dividing the total number of events to total follow up time in person-days. Kaplan Meier survival analysis was used to estimate the cumulative risk of mortality at a specific point in time and the overall median time to death of the asphyxiated neonates. The log-rank test was used to assess the statistically significant median time to death difference between categories of covariates at p<0.05.

Multicollinearity was assessed between independent variables by using a variance inflation factor (VIF). In this study the maximum VIF value was 1.654 and the mean VIF was 1.296. Therefore, there was no multicollinearity among independent variables, and we can make valid interpretation of the results of the final multivariate regression model.

Testing the proportional hazard assumption (PHA) is vital to interpret and to use fitting proportional hazard models and accept multivariate analysis results. In this study we used the Schoenfeld’s residuals test and log-log plot of survival or test of parallel lines to test the PHA. The Schoenfeld’s residuals PHA test was run for the individual covariates and global test was used after running the final model. Accordingly, the p-value for the global test is 0.990 (X^2^=7.02), which is greater than 0.05. Therefore, in this study the PHA was not violated. Graphical representations of the PHA test for some of the covariates included in the final regression model are shown in Fig 1.

**Fig 1.**
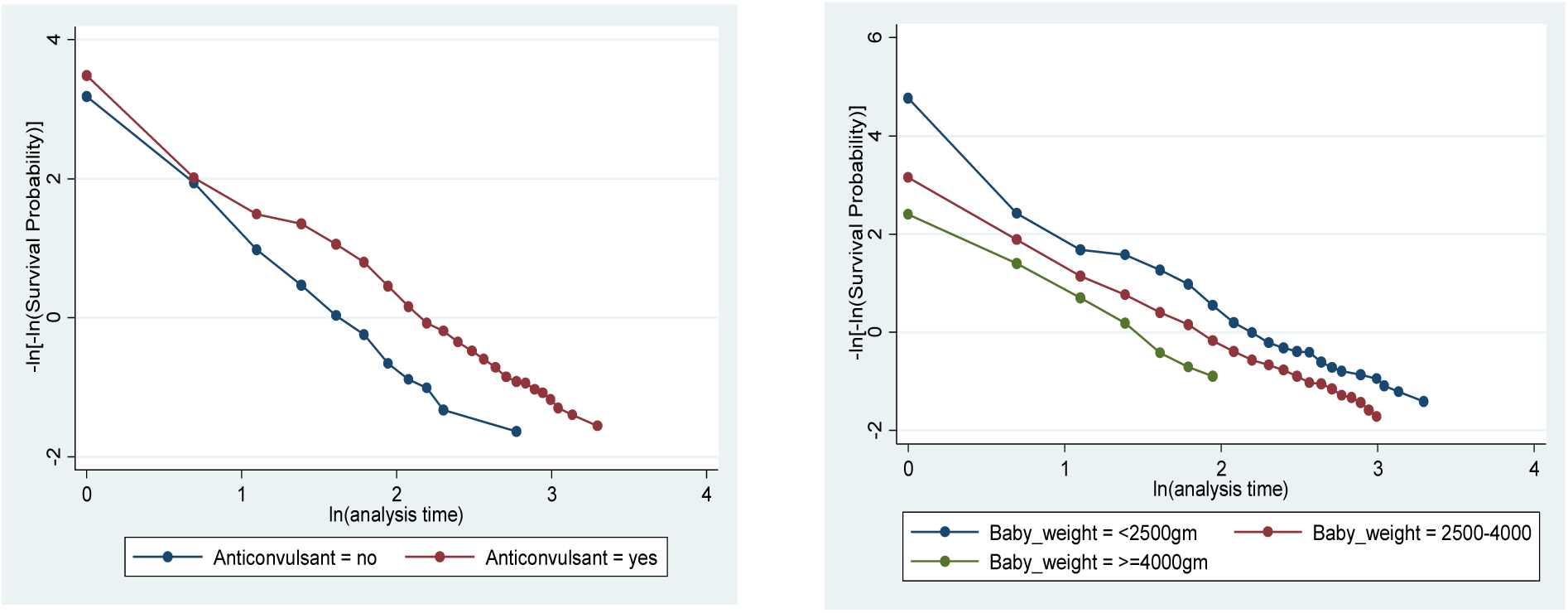
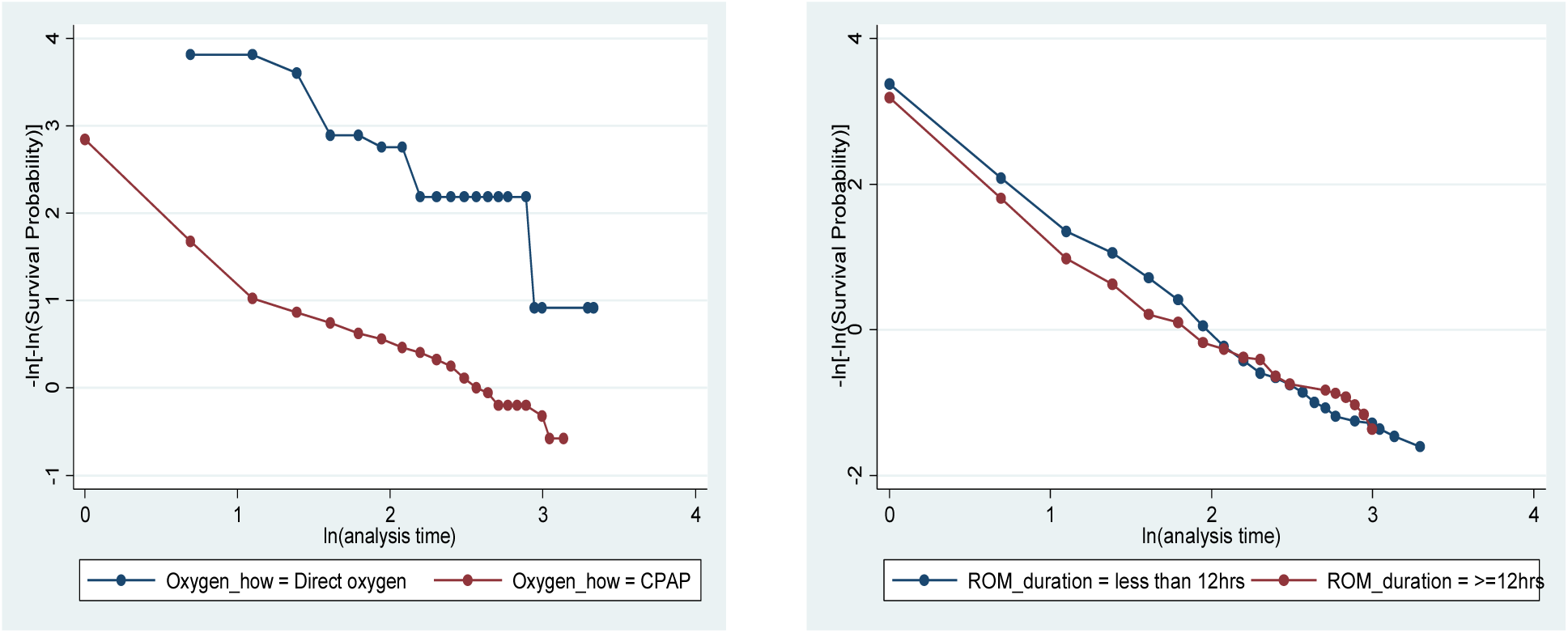
Test of proportional hazard assumption graphically by using test of parallel lines for treatment with anticonvulsant, birth weight, method of oxygen administration, and duration of ROM.

Model comparison was made between Cox, Weibull, Exponential, and Gompertz regression models using Akaki’s Information Criteria (AIC) and log-likelihood to determine which model was the best fit for the given data set. The model with the lowest AIC was taken as the best fit. Therefore, as shown on

**Table 2** the Weibull regression with an AIC of 280.75 was the model that suited the data.

**Table 2.** Summary of model comparison among the semi-parametric (Cox proportional hazard model) and parametric regression models using AIC, BIC, and Log-likelihood testes.

| Model | AIC | BIC | Log-likelihood |
| --- | --- | --- | --- |
| Cox | 538.57 | 601.52 | -251.28 |
| Weibull | 280.75 | 350.70 | -120.38 |
| Exponential | 287.46 | 353.91 | -124.73 |
| Gompertz | 285.90 | 355.84 | -122.95 |

Cox-Snell residuals test is used to assess the overall goodness of fit of our model. In this study, as seen on **Fig 2**, the hazard function follows the 45-degrees line closer to the baseline. Therefore, it showed that our multivariate Weibull-regression model is fitted for the analysis and interpretation of our data.

**Fig 2.**
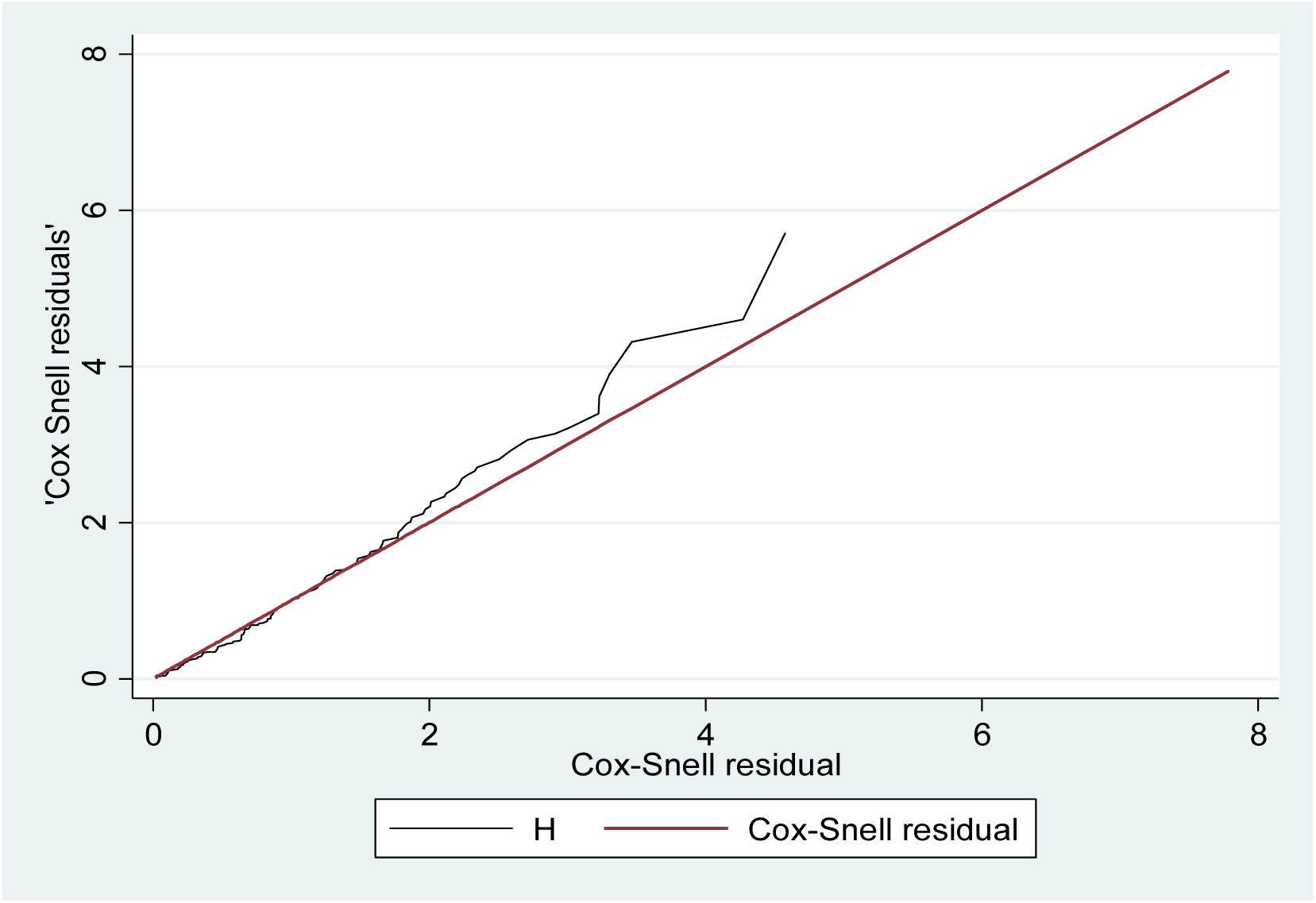
Cox-Snell residuals graph of the final Weibull regression model in the study of the time to death and its predictors among asphyxiated neonates admitted to WSUCSH, Southern Ethiopia; 2024.

Finally, to identify independent predictors of survival of asphyxiated neonates, Weibull proportional hazards regression model was used. First Bi-variable Weibull regression model was fitted for each explanatory variable, then, those variables having a P-value<0.2 in bivariate analysis, non-collinear, and those which doesn’t violate the PH assumption were entered in the final multivariate analysis. Hazard ratio with 95% confidence interval and p-values (<0.05) was used to measure the strength of association and to identify statistically significant predictors.

### Ethics approval and consent to participate

An ethical clearance letter was obtained from Wolaita Sodo University Collage of Health Sciences and Medicine (CHSM) institutional review board. Then support letter was obtained from the CHSM and was submitted to the hospital’s chief clinical director office to grant permission to access the medical records of the study subjects. All information collected from patients’ chart were kept under strict confidentiality and not disclosed to any person other than principal investigator and data collectors. Care was taken not to include personal identifiers such as names of the participants.

## Results

Four medical records with incomplete data were excluded from the initial sample of 408, leaving 404 asphyxiated neonates for final analysis. These neonates were followed for a total of 2,889 person-days (a unit combining the number of patients and the duration of observation). During this period, 89 (22.0%) of the neonates passed away (event of interest), while the remaining 315 (78.0%) censored. Fig 3 illustrates the overall outcomes for these neonates.

**Fig 3.**
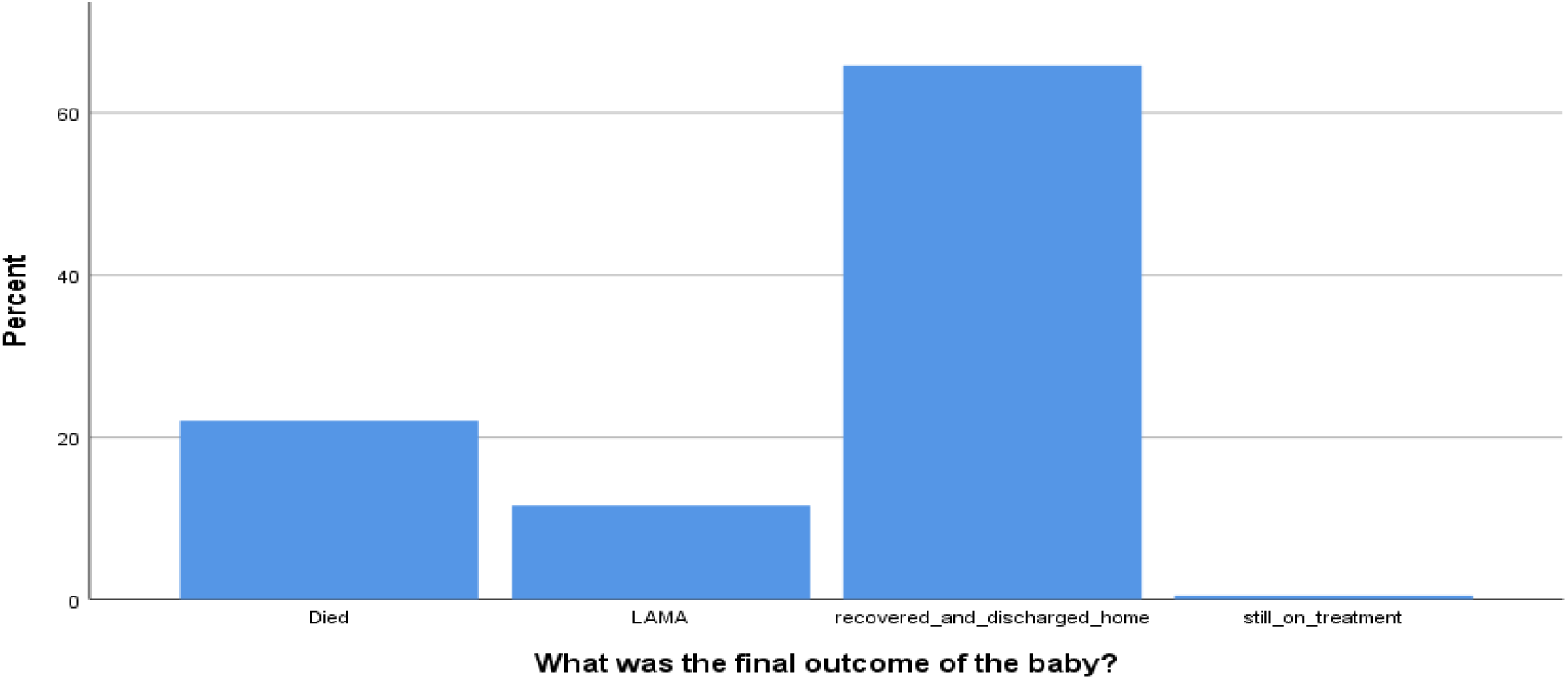
The overall outcomes of asphyxiated neonates who were admitted at the NICU of WSUCSH, Southern Ethiopia; 2024.

### Sociodemographic & obstetric characteristics of mothers of study participants

The mothers of the asphyxiated neonates were primarily middle aged adults, with most (80.4%) falling between 21 and 34 years old (mean age: 26 years, SD: 4.8). A majority (55.7%) resided in rural areas. Regarding parity, multiparous mothers (those with two or more previous pregnancies) were the most common group (59.7%). Prenatal care and delivery location showed positive trends. Over 70% of the mothers had at least one antenatal care (ANC) visit, and a similar proportion delivered at Wolaita Sodo University Comprehensive Specialized Hospital (WSUCSH) itself (inborn deliveries). The most frequent delivery mode was spontaneous vertex delivery (SVD) at 47%, followed by cesarean section (C-section) at 35.1% (Table 3).

**Table 3.** Sociodemographic and obstetrics characteristics of mothers of asphyxiated neonates who were admitted to WSUCSH, Southern Ethiopia; 2024.

| S. No. | Variables | Response | Total freq.(percent) | Survival status |  |
| --- | --- | --- | --- | --- | --- |
|  |  |  |  | Died (n/%) | Censored (n/%) |
| 1 | Age of the mother (n=404) | ≤20 years | 52 (12.9) | 8(15.4) | 44(84.6) |
|  |  | 21-34 years | 325 (80.4) | 74(22.8) | 251(77.2) |
|  |  | ≥35 years | 27 (6.7) | 7(25.9) | 20(74.1) |
| 2 | Residence of the mother (n=404) | Urban | 179(44.3) | 38(21.2) | 141(78.8) |
|  |  | Rural | 225(55.7) | 51(22.7) | 174(77.3) |
| 3 | Parity (n=404) | Primiparous | 163(40.3) | 32(19.6) | 131(80.4) |
|  |  | Multiparous | 241(59.7) | 57(23.7) | 184(76.3) |
| 4 | ANC follow-up (n=400) | Yes | 289(71.5) | 68(23.5) | 221(76.5) |
|  |  | No | 111(27.5) | 21(18.9) | 90(81.1) |
| 5 | Place of delivery (n=404) | In born (WSUCSH) | 292(72.3) | 68(23.3) | 224(76.7) |
|  |  | Out born | 112(27.7) | 21(18.8) | 91(81.3) |
| 6 | Mode of delivery (n=404) | SVD | 190(47) | 33(17.4) | 157(82.6) |
|  |  | Cesarean section | 142(35.1) | 41(28.9) | 101(71.1) |
|  |  | Instrumental and assisted breach delivery | 72(17.9) | 15(20.8) | 57(79.2) |
| 7 | Duration of labor (n=400) | Less than 24 hours | 337(84.3) | 74(22.0) | 263(78.0) |
|  |  | Greater than 24 hours | 63(15.7) | 15(23.8) | 48(76.2) |
| 8 | Duration of ROM (n=398) | Less than 12 hours | 299(75.1) | 58(19.4) | 241(80.6) |
|  |  | Greater than 12 hours | 99(24.9) | 31(31.3) | 68(68.7) |
| 9 | Type of pregnancy (n=404) | Singleton | 370(91.6) | 83(22.4) | 287(77.6) |
|  |  | Twin and above | 34(8.4) | 6(17.6) | 28(82.4) |
| 10 | Amniotic fluid (n=367) | Clear | 103(28.1) | 4(3.9) | 99(96.1) |
|  |  | Meconium stained | 264(71.9) | 72(27.3) | 192(72.7) |

### Neonatal characteristics

Slightly over half (56.2%) of the neonates diagnosed with perinatal asphyxia were male. The majority were born at term gestation (70%) and with normal birth weight (65.1%). The average birth weight was 2,959 grams (g) with a SD of 794g. Notably, two-thirds of these babies did not cry immediately after delivery, and most (74.2%) had additional comorbidities besides PNA (Table 4).

**Table 4.**
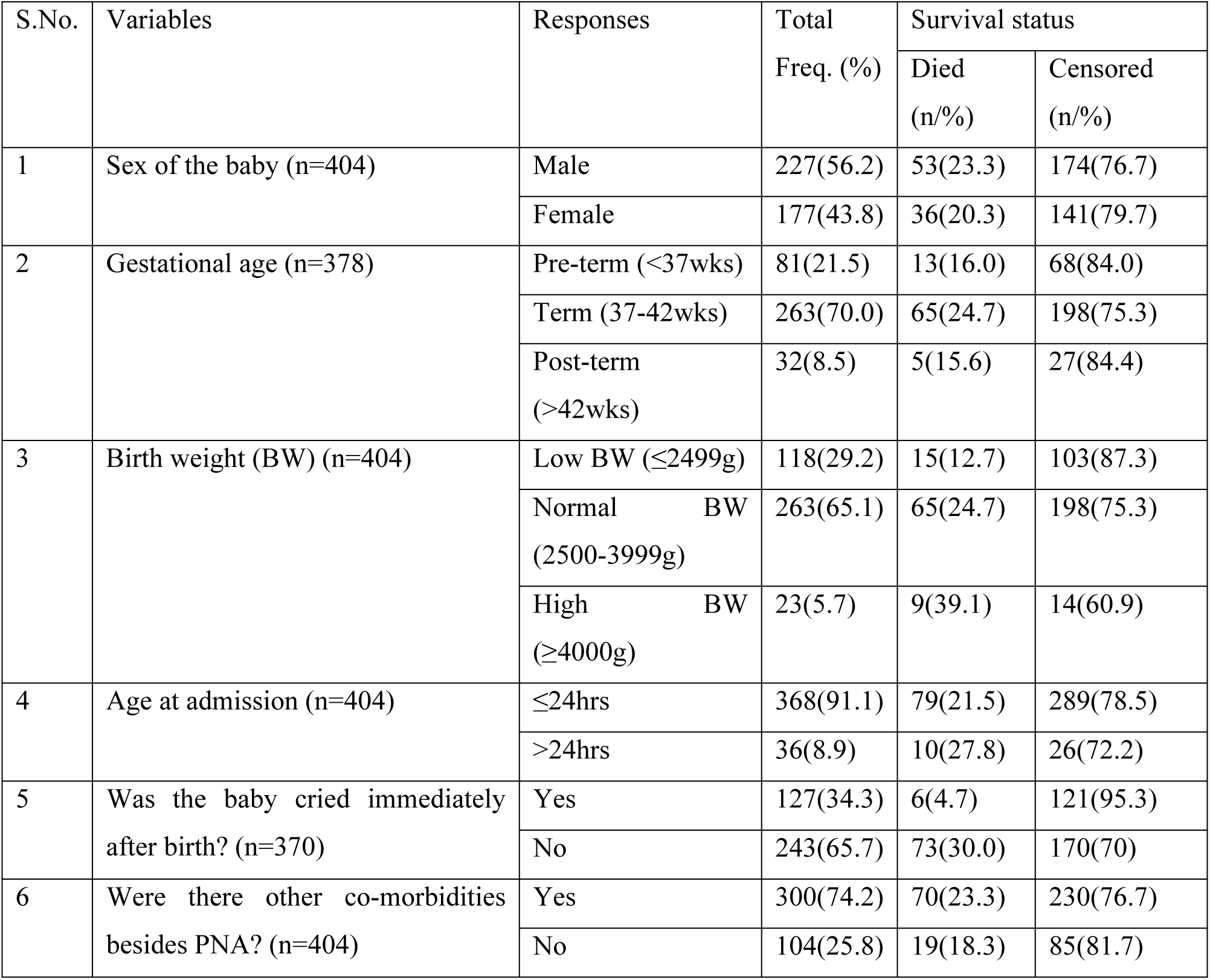
Neonatal characteristics of asphyxiated neonates who were admitted to the NICU of WSUCSH, Southern Ethiopia; 2024.

| S.No. | Variables | Responses | Total<br>Freq. (%) | Survival status |  |
| --- | --- | --- | --- | --- | --- |
|  |  |  |  | Died<br>(n/%) | Censored<br>(n/%) |
| 1 | Sex of the baby (n=404) | Male | 227(56.2) | 53(23.3) | 174(76.7) |
|  |  | Female | 177(43.8) | 36(20.3) | 141(79.7) |
| 2 | Gestational age (n=378) | Pre-term (<37wks) | 81(21.5) | 13(16.0) | 68(84.0) |
|  |  | Term (37-42wks) | 263(70.0) | 65(24.7) | 198(75.3) |
|  |  | Post-term (>42wks) | 32(8.5) | 5(15.6) | 27(84.4) |
| 3 | Birth weight (BW) (n=404) | Low BW (≤2499g) | 118(29.2) | 15(12.7) | 103(87.3) |
|  |  | Normal BW (2500-3999g) | 263(65.1) | 65(24.7) | 198(75.3) |
| | | High BW (≥4000g) | 23(5.7) | 9(39.1) | 14(60.9) |
| 4 | Age at admission (n=404) | ≤24hrs | 368(91.1) | 79(21.5) | 289(78.5) |
|  |  | >24hrs | 36(8.9) | 10(27.8) | 26(72.2) |
| 5 | Was the baby cried immediately after birth? (n=370) | Yes | 127(34.3) | 6(4.7) | 121(95.3) |
|  |  | No | 243(65.7) | 73(30.0) | 170(70) |
| 6 | Were there other co-morbidities besides PNA? (n=404) | Yes | 300(74.2) | 70(23.3) | 230(76.7) |
|  |  | No | 104(25.8) | 19(18.3) | 85(81.7) |

### Clinical Findings and Management

Upon admission, most asphyxiated neonates had low Apgar scores (below 7) at all three measured time points (1st, 5th, and 10th minute). Diagnoses included hypoxic-ischemic encephalopathy (HIE) with varying severity stages (30% stage I, 47.5% stage II, and 22.5% stage III). During their hospital stay, the most frequent complications were hospital-acquired infections (HAIs) (54.3%), seizures (33.3%), and hypoglycemia (20.4%) (refer to **S1 Table** for details).

Over 76% of the asphyxiated neonates received oropharyngeal suctioning and resuscitation. Resuscitation methods varied, with 60% requiring only bag-and-mask ventilation, while the remaining 40% needed advanced techniques like chest compressions, intubation, and adrenaline. Most neonates received oxygen therapy (88.4%), antibiotics (91.8%), intravenous fluids (94.3%), and anticonvulsants (57.4%) as part of their PNA management (refer to **S2 Table** for details).

### Incidence of mortality and median time to death

According to the study, the overall incidence density of mortality was 30.8 per 1000 person days (95%CI: 25.0-37.9). The estimated cumulative proportion of surviving on days 1, 7, 14, and 21 are 0.98, 0.817, 0.648 and 0.39 respectively. Sixty-four (71.91%), of the deaths of the neonates occurred during the first 7 days of follow-up period. The median time to death was 20 days (95% CI: 16.0, 23.9) (Fig 4).

**Fig 4.**
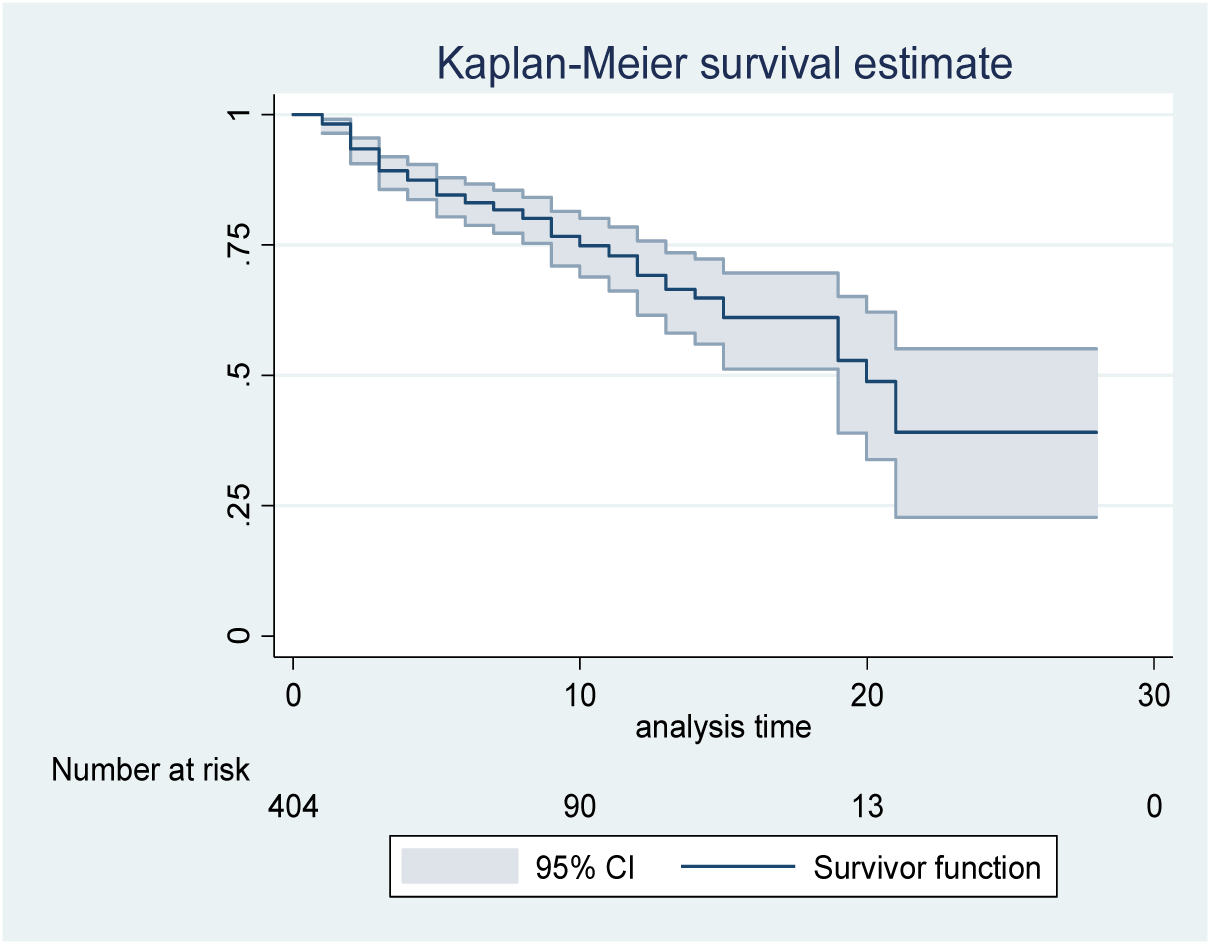
Kaplan Meier survival plot of neonates admitted with perinatal asphyxia to WSUCSH, Southern Ethiopia; 2024.

As shown on Fig 4, the overall Kaplan Meier survival curve depicts the probability of survival decreases as the follow up time increases. During the 1^st^ day of hospital stay the probability of survival was 98%, and on the 7^th^ day it was 81.7%. On the 20^th^ follow up day the probability of survival was 50% which is the overall median survival day.

### Time to death difference among categories of different covariates

Log-rank test was conducted to assess the existence of any significant difference in survival time among categories of different explanatory variables. Accordingly, a statistically significant difference in the survival time among categories of stage of HIE, duration of rupture of membrane (ROM), weight of the baby, fifth-minute Apgar score, respiratory rate at admission, presence of ANC follow up, condition of amniotic fluid, mode of delivery, and gestational age at delivery was observed at p<0.05. (Fig 5)

**Fig 5.**
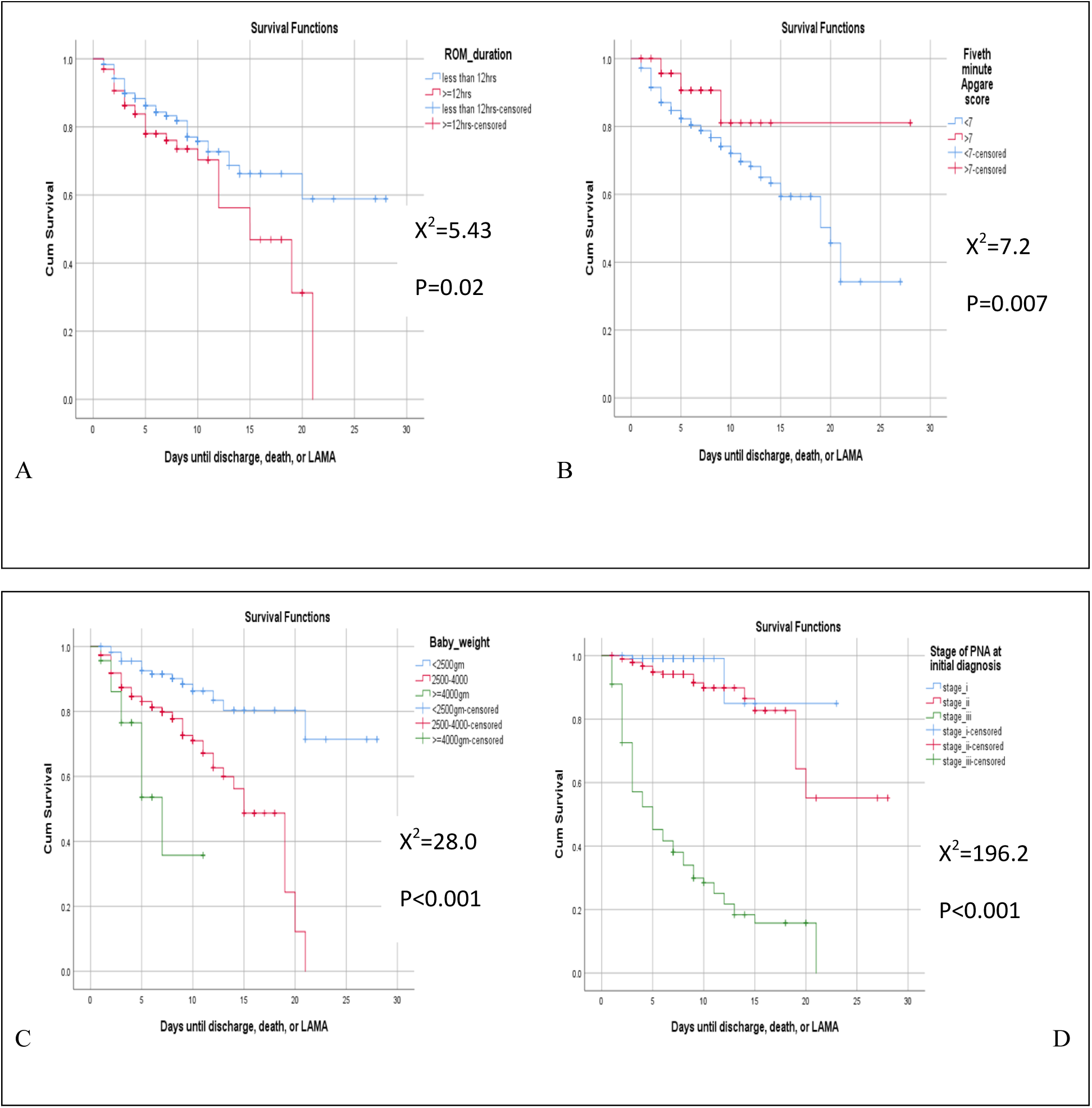
The KM survival curves that compare time to death with different categories of (A) ROM duration, (B) fifth-minute Apgar score, (C) birth weight, and (D) stage of HIE at admission among asphyxiated babies who were admitted to NICU of WSUCSH.

As shown on Fig 5-A neonates who are delivered after a prolonged duration of rupture of membrane (ROM) (red line) has lower survival as compared with those with normal duration of ROM (blue line) and the difference is statistically significant at p<0.05. Similarly, those neonates with fifth-minute Apgar score of greater than 7 (Fig 5-B (red line) has a better survival as compared with those who has Apgar score less than 7 at the fifth-minute (blue line) and the difference is statistically significant at p<0.05. On log-rank test as shown on Fig 5-C, babies with low birth weight (blue line) has higher survival than those with normal (red) and high birth weight (green line). According to this study the median time to death among neonates with stage-III HIE is much shorter 5 days (95% CI: 2.95-7.05) or they have a shorter survival time as compared to the overall median survival time (20days). This difference was statistically significant at p-value < 0.0001 (Fig 5-D).

### Predictors of time to death among asphyxiated neonates

The relationship between the independent and outcome variables was analyzed using Weibull proportional hazard regression model. According to this study, on multi-variable Weibull regression analysis, the predictors of time to death among asphyxiated neonates at p<0.05 were: 5^th^-minute Apgar score greater than seven, birth weight greater than 4000gm, meconium stained amniotic fluid during delivery, oxygen saturation level at admission, oxygen administration with Continuous Positive Airway Pressure (CPAP), resuscitation with chest compression, and need for anticonvulsant.

Keeping other variables constant, neonates who were delivered with meconium stained amniotic fluid were 14 times more likely to die (AHR=14.40; 95%CI:3.02-68.57) compared with those who delivered with clear amniotic fluid. Neonates with a fifth-minute Apgar score of less than 7 has 3.4 times higher hazard of death (AHR=3.42; 95%CI: 1.43-8.18) compared to those with a normal Apgar score. Similarly, neonates with high birth weight (BW>4000gms) are 4.7 times more likely to die faster (AHR=4.74; 95%CI: 1.53-14.71) than those with normal birth weight. At any point in time throughout the follow-up period, a unite increase in the admission oxygen saturation level results in a 5% decrease in the hazard of death due to PNA (AHR=0.95; 95%CI: 0.93-0.97). Three management related variables, namely; treatment with anticonvulsants, method of oxygen administration, and resuscitation method were significant predictors of time to death after PNA in the study. Accordingly, neonates who were treated with anticonvulsant have 6 times higher hazard of death (AHR=6.44; 95%CI: 1.41-29.41) as compared with those who didn’t needed treatment with anticonvulsant. Neonates who took oxygen through CPAP are 2.7 times more likely to die (AHR=2.790; 95%CI; 1.19-6.53) compared to those who took direct oxygen. Moreover, neonates who needed chest compression during resuscitation are 5 times more likely to die than those who were resuscitated using bag and mask only (AHR=5.09; 95%CI: 2.34-11.06) (**Table 5**).

**Table 5.** Bi-variable and Multi-variable Weibull regression analysis for the predictors of the time to death among neonates who were admitted with PNA at WSUCSH; Southern Ethiopia, 2024.

| Covariates | CHR (95%CI) | AHR (95% CI) | P- value |
| --- | --- | --- | --- |
| ROM duration (Ref. less than 12hrs) |  |  |  |
| Greater than 12 hours | 1.69(1.09-2.62) | 0.57(0.22-1.48) | 0.254 |
| Condition of amniotic fluid during delivery (Ref. Clear) |  |  |  |
| MSAF | 7.85(2.86-21.50) | 14.40(3.02-68.57) | **0.001\*** |
| Mode of delivery (Ref. Spontaneous vertex delivery (SVD)) |  |  |  |
| Cesarean section | 1.71(1.08- 2.70) | 0.70 (0.30-1.60) | 0.403 |
| Instrumental and assisted breech delivery | 1.06(0.57- 1.9) | 1.37 (0.50- 3.71) | 0.532 |
| Apgar score-5<sup>th</sup> minute (Ref. Greater than 7) |  |  |  |
| Less than 7 | 2.478(1.24- 4.95) | 3.42(1.43-8.18) | **0.006\*** |
| Birth weight in grams (Ref. 2500-3999) |  |  |  |
| Less than 2500 | 0.30(.17- .53) | 0.34(.13-.91) | 0.053 |
| >= 4000 | 2.64(1.31-5.35) | 4.74(1.53-14.71) | **0.007\*** |
| Pulse rate at admission | 0.99(.98-1.00) | 1.00(.99-1.02) | 0.098 |
| Temperature level at admission | 0.65(.53-.79) | 1.16(.87-1.56) | 0.301 |
| Oxygen saturation level at admission | 0.913(.89- .93) | 0.95(0.93-0.97) | **0.005\*** |
| RBS level at admission | 0.99(.98-1.00) | 1.00(.99-1.00) | 0.733 |
| Medical complications that occurred during hospital stay/ Acute kidney injury (AKI) (Ref. No) |  |  |  |
| Yes | 1.798(1.13- 2.87) | .93(.39-2.22) | 0.881 |
| Medical complications that occurred during hospital stay/ seizure (Ref. No) |  |  |  |
| Yes | 2.656(1.724- 4.094) | 1.05(.56-1.98) | 0.869 |
| Was the baby given anticonvulsant? (Ref. No) |  |  |  |
| Yes | 6.84(2.96- 15.78) | 6.44 (1.41-29.41) | **0.016\*** |
| Oropharyngeal suctioning done? (Ref. NO) |  |  |  |
| Yes | 4.98(1.82-13.60) | .35(.06-2.09) | 0.255 |
| Method of oxygen administration (Ref. Direct method) |  |  |  |
| CPAP | 7.83(4.61- 13.32) | 2.790 (1.19-6.53) | **0.018\*** |
| Method of resuscitation (Ref. Bag and mask only) |  |  |  |
| Chest compression | 4.73(2.97- 7.54) | 5.09(2.34-11.06) | **0.000\*** |
| Intubation and adrenalin | 3.90(2.15- 7.09) | 1.22(.46-3.22) | 0.679 |

*\*statistically significant association ( $p < 0.05$ )*
*AHR=adjusted hazard ratio, CHR=crude hazard ratio, CI= confidence interval, ROM=rupture of membrane, MSAF=meconium Stained Amniotic Fluid, CPAP=continuous positive airway pressure*

## Discussion

This retrospective follow-up study determined the time to death and identified its predictors among 404 asphyxiated neonates admitted to the NICU at WSUCSH. Among these neonates, 22% (89) died during the follow-up period. The incidence density of mortality due to PNA in our study was 30.8 per 1000 person days (95%CI: 25.0-37.9), which is comparable to findings from other tertiary hospitals in Ethiopia (Jimma: 29/1000 (95% CI: 23.20-36.01), Addis Ababa: 32/1000, and West Oromia: 38.86/1000 (95% CI: 33.85–44.60)) [19, 20, 24]. These similarities highlight the significant burden of PNA-related mortality across the country. While variations in sample size and study period might explain slight differences in incidence density, our findings suggest a persistent challenge. Furthermore, considering the national neonatal mortality rate of 33 per 1000 live births reported in the 2019 mini-EDHS [15], our institutional data might underestimate the true national picture. Therefore, a large-scale national study is crucial to obtain a more comprehensive understanding of PNA-related mortality in Ethiopia.

The incidence of mortality due to PNA in our study (30.8 per 1000 person days) falls between the findings reported from other regions in Ethiopia. Compared to studies in Dessie (53/1000) and North-West Ethiopia (53.49/1000) [18, 26], our results suggest a potentially lower mortality burden. This difference might be attributed to variations in study setting and resources: Our study was conducted at WSUCSH, a tertiary hospital with potentially better access to human resources and advanced equipment compared to the facilities in the aforementioned studies. The other reason might be due to variations in temporal trends in treatment: Our data spans five years (more recent than the three-year periods in the compared studies). This might reflect improvements in neonatal care practices over time.

However, our findings also show a higher mortality rate compared to studies in South-West Ethiopia (9.1/1000) and Nigeria (12.8/1000) [25, 27]. Here, two potential explanations can be given: the first explanation is difference in study design and data Source: Our retrospective design utilizing medical records might overestimated mortality compared to the prospective cohort studies used in the other studies, which likely followed patients more closely. The second explanation is difference in study setting and population characteristics: The South-West Ethiopian study involved multiple facilities, potentially representing a broader population with varying risk factors. Additionally, geographical and sociodemographic differences, including healthcare access and utilization, might contribute to the observed discrepancy with the Nigerian study. Therefore, a prospective cohort study should be done in the current study facility to have a better picture of the incidence of mortality due to PNA and for a better comparison with other studies.

The median time to death in our study was 20 days, similar to the findings from Jimma (20 days) [20]. However, this is somewhat longer than the median survival times reported in West Oromia and North West Ethiopia (15 days) [24, 26]. A more striking difference emerges when compared to studies in Dessie and South West Ethiopia, which documented a median time to death of only 8 days [18, 25]. Several potential explanations for this variation warrant further investigation. These might include study design difference, as the study conducted in South West Ethiopia is a prospective cohort and followed the actual patients. Even though, the study conducted in Dessie is a retrospective cohort, there might be differences in the characteristics of the study population, differences in the severity of asphyxia at presentation and variations in treatment protocols. Additionally, slight variations in how time to death was defined or ascertained in different studies might play a significant role; for example, in the South West Ethiopian study they used the KM failure curve to report the median time to death. The retrospective nature of our study also limits our ability to definitively pinpoint the reasons for this variation. Future multicenter studies with standardized data collection methods are needed to explore the factors influencing survival time after PNA and to understand the observed differences between different facilities across the country in median time to death.

Our study identified the presence of meconium-stained amniotic fluid (MSAF) during delivery as a significant predictor of time to death from PNA. Newborns delivered with MSAF were 14 times more likely to die (AHR = 14.40; 95% CI: 3.02-68.57) compared to those delivered with clear fluid. This increased mortality risk is likely due to the association between MSAF and adverse birth outcomes, including perinatal asphyxia, neonatal sepsis, and meconium aspiration syndrome (MAS) [28]. A study in Northwest Ethiopia supports this finding, reporting a higher incidence of perinatal asphyxia among newborns born with MSAF (13.8%) compared to those without (7.6%) [26]. These findings highlight the importance of heightened awareness among labor and delivery staff regarding the potential complications associated with MSAF. Prompt delivery and coordinated resuscitation strategies should be implemented upon MSAF diagnosis to minimize the risk of PNA-associated neonatal mortality.

Our analysis revealed that neonates with a low fifth-minute Apgar score (less than 7) were 3.4 times more likely to die (AHR= 3.42; 95% CI: 1.43-8.18) compared to those with a normal score. This finding aligns with research conducted in West Oromia [24], highlighting the established link between low Apgar scores and adverse outcomes. As a critical indicator of a newborn’s immediate condition and potential for perinatal asphyxia, it also plays a vital role in guiding resuscitation efforts [29]. Furthermore, large-scale studies have documented the association between low Apgar scores and long-term health risks, including cardiovascular diseases and childhood mortality [30]. Early and effective resuscitation protocols, tailored to the severity of the Apgar score, are crucial to improve outcomes for neonates experiencing birth asphyxia.

Our study revealed a significant association between high birth weight (macrosomia, >4000 grams) and a 4.7-fold increased hazard of death from PNA (AHR=4.74; 95% CI: 1.53-14.71). This finding aligns with research conducted in Dessie and Addis Ababa [18, 19], highlighting the potential risks associated with macrosomia. A large European study further supports this link, demonstrating a 4-fold increase in perinatal asphyxia risk among macrosomic babies compared to those with normal birth weight [31]. These observations emphasize the need for increased vigilance and potentially modified delivery strategies for pregnancies identified as high-risk for macrosomia. Early identification and close monitoring of these pregnancies may allow for interventions to optimize fetal well-being and potentially reduce the risk of PNA and associated mortality.

Our study found that a one-unit increase in admission oxygen saturation was associated with a 5% decrease in the hazard of death from PNA (AHR= 0.95; 95% CI: 0.93-0.98). This aligns with research from Southeast Nigeria [27], highlighting the critical role of adequate oxygenation in improving outcomes for neonates with PNA. The link between oxygen saturation and mortality risk is further supported by a study in Northwest Ethiopia, which reported a significantly higher hazard of death among neonates with lower oxygen saturation levels [26]. These findings underscore the importance of prompt oxygen therapy and close monitoring of oxygen saturation for asphyxiated neonates to prevent hypoxia and its detrimental consequences, such as hypoxic encephalopathy.

Our study identified several key management practices that significantly influence time to death after PNA. These include treatment with anticonvulsants, oxygen administration method, and resuscitation technique. Neonates receiving anticonvulsants had a 6-fold increased hazard of death (AHR = 6.44; 95% CI: 1.41-29.41) compared to those who did not. This association likely reflects the underlying severity of their condition, as anticonvulsants are typically used for neonates with seizures or severe PNA (HIE stage II and III), conditions known to be major mortality predictors [18, 19, 22]. These findings highlight the importance of accurate diagnosis and targeted treatment strategies for PNA patients.

Furthermore, the method of oxygen administration impacted mortality risk. Neonates receiving oxygen via CPAP were 2.7 times more likely to die (AHR = 2.790; 95% CI: 1.19-6.53) compared to those receiving direct oxygen. Similarly, neonates requiring chest compressions during resuscitation had a 5-fold increased risk of death (AHR = 5.09; 95% CI: 2.34-11.06) compared to those resuscitated with bag-and-mask only. These observations emphasize the crucial role of timely and appropriate respiratory support and resuscitation techniques tailored to the severity of the neonate’s condition. Successful neonatal resuscitation efforts depend on critical actions that must occur in rapid succession; hence, the availability of resuscitation equipment, drugs, and experienced personnel are critical to maximize the chances of survival [32]. Investing in equipping neonatal units with essential resuscitation equipment and ensuring the availability of skilled personnel are critical steps to improve outcomes for asphyxiated neonates. A study in Ethiopia revealed that only half of healthcare facilities were adequately equipped for neonatal resuscitation [33]. Addressing these resource limitations is essential to optimize survival rates.

## Limitations of the study

This study has some limitations inherent to its facility based retrospective design using secondary data sources. Missing maternal data on socioeconomic factors (educational status, occupation, etc.) could potentially influence neonatal mortality but were unavailable from medical records. Additionally, the study design limited our control over the quality of some measurements and classifications. For example, we relied on physician-assigned stages of PNA/HIE, which might introduce misclassification bias. Generalizability of the findings is limited due to the study’s facility-based nature. The results may not be directly applicable to facilities with less equipped NICUs, such as health centers and private clinics. Additionally, neonates who were delivered at home and not admitted to the hospital could not be included in the study, potentially biasing the sample towards more severe cases.

## Conclusion

This study identified a high mortality rate and prolonged hospitalization times among neonates with perinatal asphyxia (PNA) admitted to the WSUCSH NICU. It also pinpointed factors influencing survival, including birth complications, high birth weight, low oxygen levels, and specific treatment approaches. Based on these findings, the health facilities are recommended to implement PNA risk assessment protocols, standardized delivery and resuscitation protocols, and invest in essential equipment and staff training on resuscitation techniques. The ministry of health and various stakeholders are also recommended to collaborate to update national clinical guidelines based on these findings, and conduct a large-scale national study to gain a more precise picture of PNA-related mortality in Ethiopia.

## Data Availability

The dataset underlying the findings reported in the manuscript will be provided as a supporting information in the submitted file.

## Acknowledgments

The authors are grateful to the data collectors and supervisors for their dedication and hard work during the data collection period. We would also like to extend our appreciation to Wolaita Sodo University Comprehensive Specialized Hospital (WSUCSH) Chief Clinical Officer, Medical Record Room and Neonatal Intensive Care Unit (NICU) staff for their cooperation throughout the study process.

## Supporting information

S1_Table.docx: Clinical and laboratory characteristics

S2_Table.docx: Management related factors

S3_File.docx: Data abstraction checklist

S4_File.xls: Dataset

## Notes

### Competing Interest Statement

The authors have declared no competing interest.

### Funding Statement

The authors received no specific funding for this work.

### Author Declarations

An ethical clearance letter was obtained from Wolaita Sodo University Collage of Health Sciences and Medicine (CHSM) institutional review board.

